# Evaluating the Efficacy and Safety of Neonatal Chyme Reinfusion Therapy: A Feasibility Study using a Novel Medical Device

**DOI:** 10.1101/2024.05.08.24306960

**Authors:** E. Ludlow, T. Harrington, R. Davidson, J. Davidson, K. Aikins, G. O’Grady, I. Bissett

## Abstract

**Objectives:** Neonatal and pediatric intestinal failure related to enterostomy is an infrequent but burdensome condition associated with substantial morbidity and mortality. This study presents the development and clinical validation of a novel device to resolve these problems, by formalizing a safe and efficient enterostomy chyme reinfusion technique.

**Methods:** A novel neonatal chyme reinfusion device was designed and manufactured (‘The Insides Neo’, The Insides Company, New Zealand), prior to validation in a feasibility study in tertiary neonatal intensive care centres. Neonates with double enterostomy were recruited and commenced on chyme reinfusion therapy using the novel device to test safety, efficacy, tolerability, and usability within nursing workflows. Device and clinical outcomes were recorded along with nursing feedback. Registered under the ANZCTR, identifier no. ACTRN12621000835842p.

**Results:** Ten neonates were recruited across two centres, with a median usage duration of 37.5 (range 12-84) days. Following initiation of therapy, rate of weight gain increased from mean 68.8 ± 37.4 to 197 ± 25.0 g/week (p=0.024). Of the 7/10 neonates on PN at commencement of therapy, 4/7 were able to wean and achieve enteral autonomy. All neonates tolerated the device with uniformly positive nursing feedback and minimal time to learn and incorporate the novel device into nursing workflows. There were no device-related adverse events.

**Conclusions:** A novel device was developed and validated to be safe and effective at performing chyme reinfusion therapy in neonates. This device is anticipated to improve the clinical care and outcomes of neonatal patients with double enterostomies.

**What is Known**

- Chyme reinfusion is widely accepted and is a standard of care in many centres who care for neonates with double enterostomy’s.
- The manual application of chyme reinfusion has led to low adoption despite the many benefits.

**What is New**

- A new device that automates the process of chyme reinfusion that overcomes many of the barriers outlined in the literature.
- Uniformly positive nursing feedback that streamlines the Neonatal Intensive Care Unit (NICU) workflow.

## Background

Intestinal failure in neonatal and paediatric populations is an infrequent but burdensome condition associated with significant morbidity and mortality. Intestinal failure is defined as an impairment in gut function, resulting in an inability to absorb macronutrients, water, and electrolytes [1,2]. Short bowel syndrome secondary to massive intestinal resection is the most common cause of paediatric intestinal failure, with up to 50% of cases due to necrotising enterocolitis (NEC), especially in premature neonates [3–6]. A small bowel double enterostomy is often performed in these cases at the time of resection, contributing to an insufficient functional gut length and high output losses via the enterostomy [3]. Studies have shown that reinfusing chyme collected from the proximal limb of the enterostomy to the distal limb is a beneficial therapeutic option in neonates [3, 7–9, 11]. These benefits include supporting nutrition and growth, normalisation of fluid balance and electrolytes, and weaning from parenteral nutrition leading to enteral autonomy, improved gut maturation, and benefits to liver function [3, 11, 13]. Studies in adults have also demonstrated that chyme reinfusion therapy (CRT) is safe and beneficial [10].

CRT has been growing in popularity due to numerous benefits, however it is a complex process, requiring nurses to dedicate considerable time to manual collection, decanting and filtration before reinfusion of chyme. In addition, manual chyme reinfusion techniques can lead to an increased prevalence of stoma appliances failing due to the need to make modifications to the stoma appliance. A systematic review completed in 2020 looked at evidence of use of CRT with double enterostomy in pediatric and neonatal groups, highlighting that CRT has been employed erratically in Neonatal Intensive Care Units (NICU) worldwide [11]. However, techniques and studies have been highly variable, with heterogenous indication criteria, methods, materials, and equipment employed, and with wide-ranging clinical outcomes [11]. Severe adverse events have been rare but have included intestinal perforation and haemorrhage [11]. Stoop et al. recently concluded in a systematic review that there is limited robust evidence for prescribing pediatric CRT, due to small study numbers and varied administration of the therapy [12]. This lack of uniformity also creates practical challenges for the health care professionals, most of whom do not consider the current practices user-friendly.

The aim of this work was to develop and validate a novel clinical device to overcome these problems, by formalizing a safe and efficient chyme reinfusion technique, while improving nursing workflows for therapy administration. This paper describes the first in-human use of the device, capturing clinical evaluation data, usability outcomes, and initial safety and efficacy data.

## Methods

### Development of novel device

This project was a collaboration between engineers and clinicians to design a chyme reinfusion device that fit with common stoma appliances and consumables available in most NICUs and maintained, if not reduced, contact with the neonate in relation to stoma care. The device needed to address the issues outlined above, to create a safe and effective tool to perform chyme reinfusion to improve patient outcomes and nursing workflows. Device design references were sourced from literature, empirical experience, and feedback from clinicians on prototypes [11,12].

### Impact of innovation

The device was named The Insides Neo (The Insides Company, New Zealand) and performs CRT as per the description in **Figure 1** and **Figure 2**. It is an easy-to-use directional flow device which connects to three standard off-the-shelf components: i) a stoma appliance; ii) an ENFit syringe; iii) and a traditional soft, flexible enteral feeding / nasogastric tube. The tube is inserted in the distal enterostomy to allow CRT when appropriate. The device is then secured at the bottom of the stoma appliance and contains a directional flow valving system to enable and facilitate CRT through a single access point. CRT can be performed either by bolus or continuous reinfusion, however, the use of a syringe driver to perform continuous is recommended to control the rate of reinfusion, thus reducing the likelihood of reflux.

**Figure 1.**
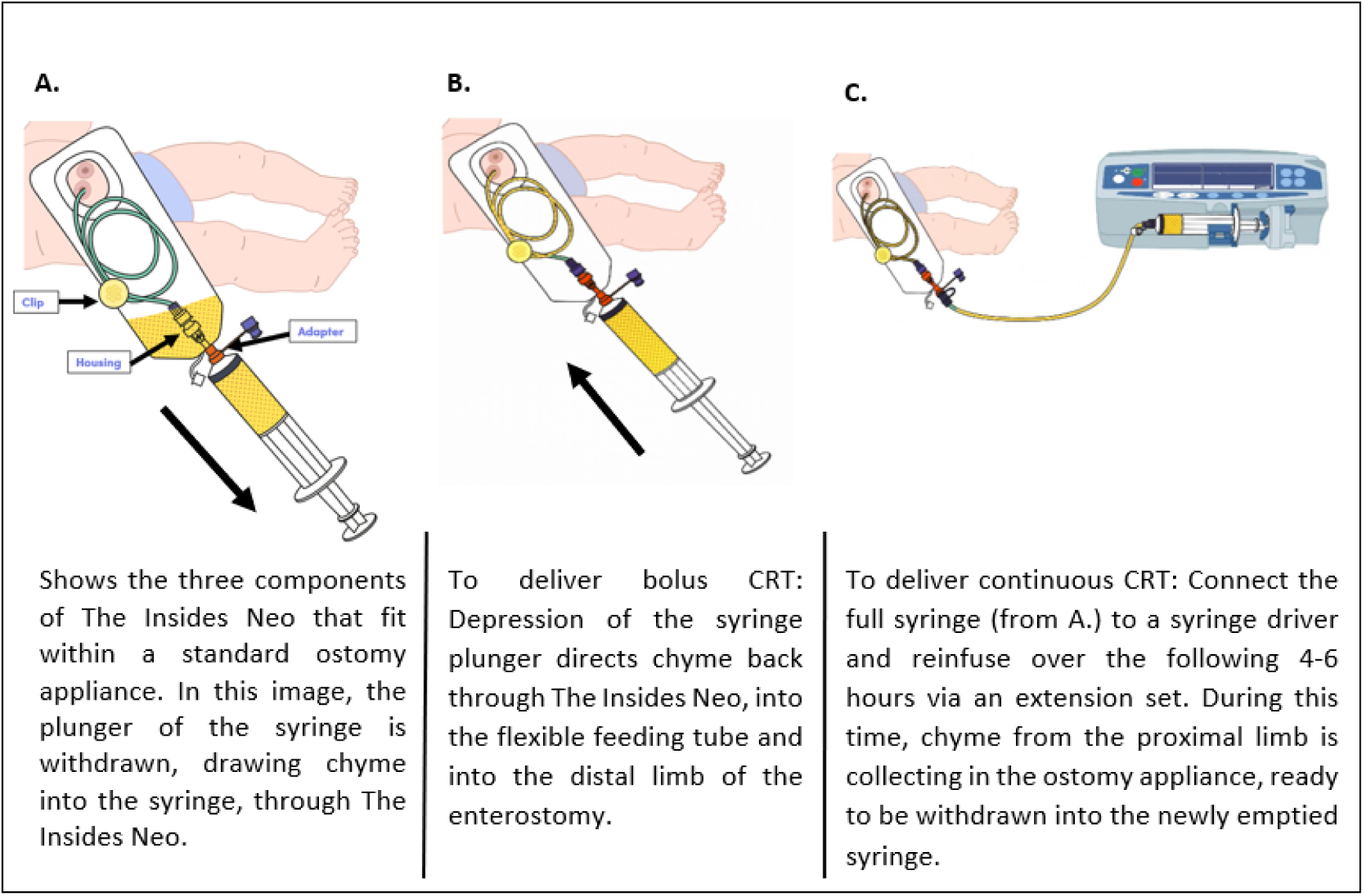
Diagram of chyme reinfusion therapy with The Insides Neo

**Figure 2.**
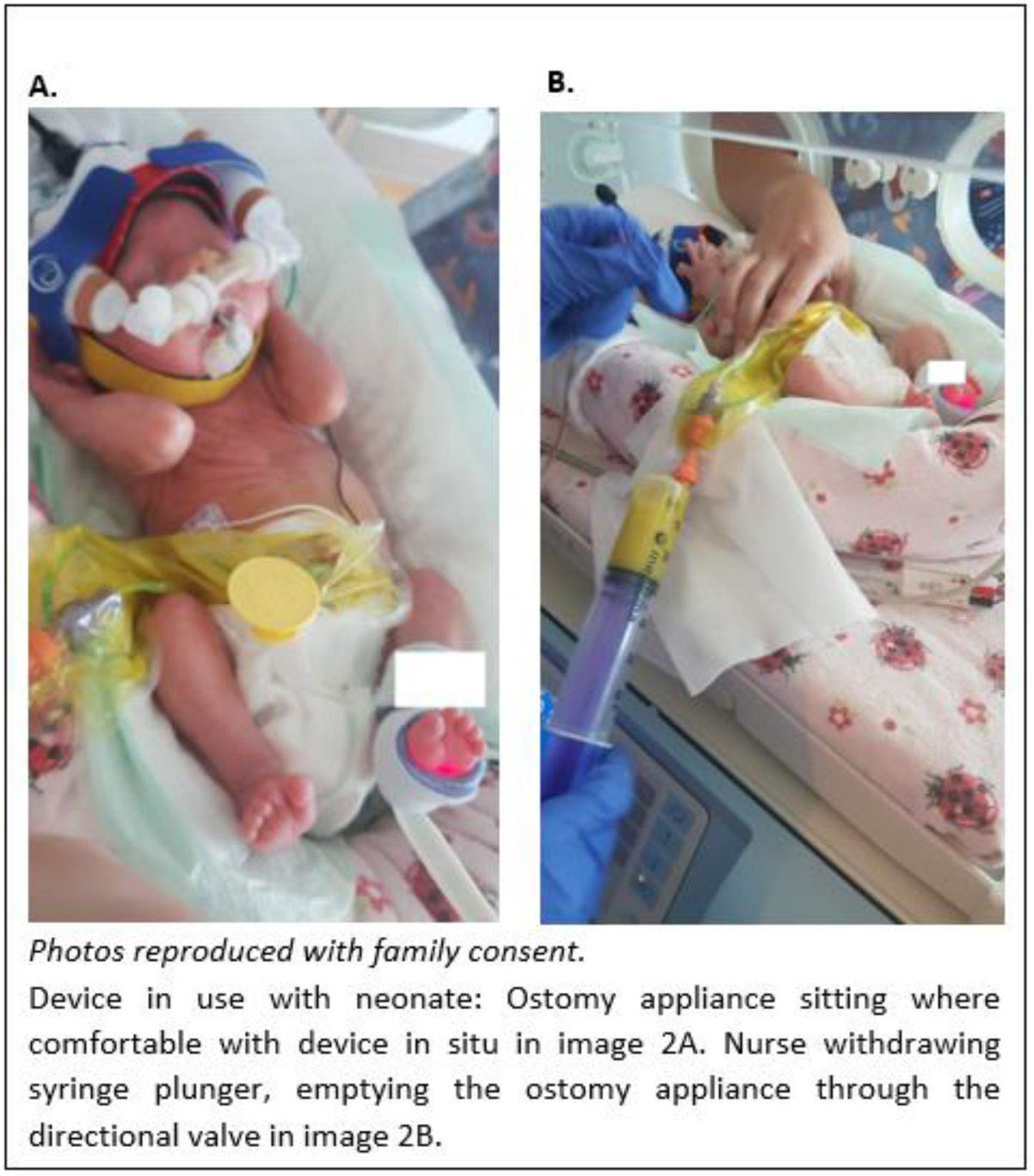
Request pictures of device in use with a Neonate from corresponding author

The device design thus overcomes several limitations that have been identified with performing manual CRT with neonates [11,12]. The Insides Neo is easy to assemble and fits with widely available stoma appliances, so no modification to the stoma appliances are required. Contact with chyme and the neonate when performing CRT is greatly reduced, together with time required, in comparison to manual CRT.

### Feasibility study

The feasibility study evaluated how the novel device fitted into nursing clinical workflows, the safety and efficacy of the device, and achieved initial clinical data including weight gain and the resumption of enteral nutrition and weaning from parenteral nutrition. This study was approved by the New Zealand Health and Disability Ethics Committee: reference 21/NTB/150. All parents of the neonates enrolled provided informed consent, including for the publication of medical photographs. The study conformed with the STROBE guidelines (**Supplementary Table 2**).

#### Primary and secondary outcomes

The primary outcomes were to validate the safety, effectiveness, and tolerability of the stoma refeeding device. Secondary outcomes were to evaluate the usability of the device in a neonatal intensive care unit (NICU) setting, and adherence to chyme reinfusion therapy protocols by the NICU staff via an anonymous feedback form. Secondary outcomes included obtaining initial clinical data from the use of the novel chyme reinfusion device, including change in weight gain rate, nutrition support therapy, and surgical reinterventions.

#### Inclusion and exclusion criteria

The following criteria were required for inclusion;

- neonates with double enterostomy or enteroatmospheric fistula with proximal and distal limbs contained in a single or two separate stoma appliances
- sufficient distal intestinal length to allow chyme or enteral nutrition
- guardians able to provide informed consent for the procedure.

The exclusion criteria included any neonate with distal limb obstruction, inability to tolerate chyme or enteral feed distally, clinical concern for ischemic gut, inability to intubate the distal limb and septic or critically unwell neonates.

#### Survey methods

A feedback form, designed for the study, was maintained at the cot-side of each neonate refeeding. Nurses caring for the patients were asked to complete this form and return it anonymously to a box. It was specified that one feedback form per nurse was to be completed over the course of their experience, with a prerequisite of having some prior experience with manual CRT to allow workflow comparisons. The feedback form consisted of 10 questions utilising a Likert scale for answers. Possible responses to each question ranged from 1 through 5 (1. Totally disagree, 2. Disagree, 3. Neither disagree nor agree, 4. Agree, 5. Totally agree).

#### Set-up and data collection

Clinical teams assessed when patients would be clinically ready to commence CRT, independent of the study investigators. The cot side nurse then intubated the distal limb with the feeding tube and applied the stoma appliance with the device assembled in the stoma appliance bung. Continuous reinfusion of the neonate’s chyme commenced and increased at a tolerable rate, as judged by the nursing team based on patient comfort and chyme reflux. Titration of nasogastric feeds and PN decisions were again made by the clinical teams, independent of study investigators. The stoma appliance (and device) was changed if it leaked, or on a schedule of up to every 4 days, informed by standard practices in adult chyme reinfusion devices [10]. Continuous reinfusion of chyme continued up to the day of enterostomy reversal, with post operative outcomes recorded by the research nurse.

Standard of care weight charts, and nutrition support records were stored in digital clinical report forms, against an anonymous study participant number. Biometric, adverse events, and side effects during therapy were recorded daily with consultation with clinical staff and monitored via weekly visits by the research nurse.

#### Analysis

Data was analysed both quantitatively and qualitatively to address the primary and secondary outcomes. Weight gain before vs after therapy was compared by the paired Student’s t-test, with a significance threshold of p<0.05.

## Results

### Neonate characteristics

Patient recruitment is detailed in **Figure 3**. Neonates were recruited from January 2022 to December 2023. Of 46 screened patients with ostomies, 10 were eligible and consented, with 36 excluded due to incorrect disease indications (n=21), transfer or death (n=9), inaccessible distal limb (n=3) and inability to fit the prescribed stoma appliance due to prolapse (n=3). Characteristics of the included neonates including disease aetiologies are outlined in **Figure 3** and **Table 1**. Of the ten neonates, six were female. All had a double barrel enterostomy formed in the small bowel with one neonate having the distal limb originating from the ascending colon. The neonates were all in a tertiary NICU setting with 1:1 nursing.

**Figure 3.**
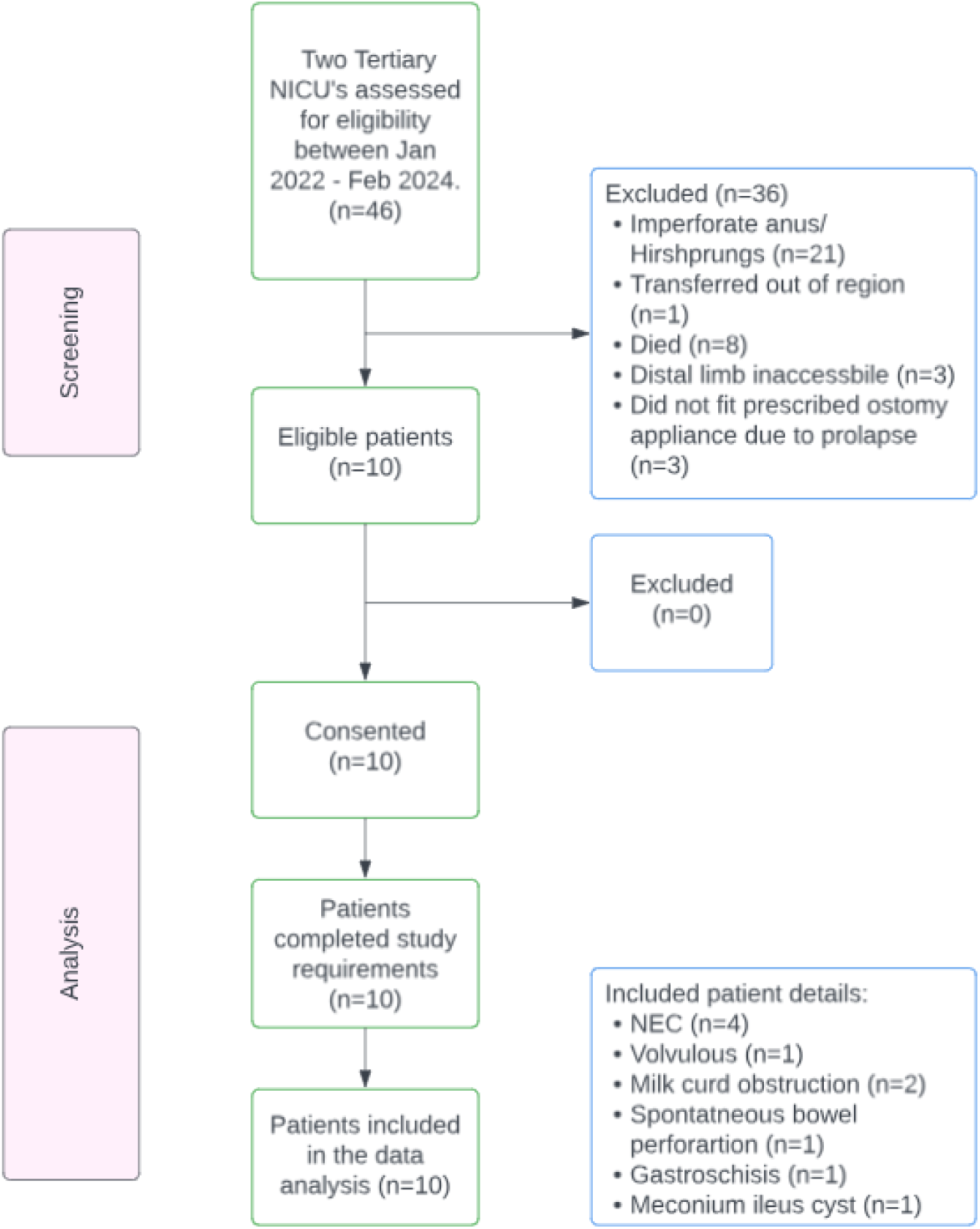
Patient recruitment flow diagram

**Table 1.** Neonate characteristics.

| Neonate no. | Sex | Ethnicity | Aetiology | Distal small bowel length (cm) | Weight at baseline prior to starting CRT (g) | Average weekly weight gain prior to CRT (g/week) |
| --- | --- | --- | --- | --- | --- | --- |
| 1. | F | NZ European | Mid gut volvulus | 40 | 1160 | 77 |
| 2. | M | Pacifica | Spontaneous bowel perforation | 10 | 1230 | 176 |
| 3. | M | Indian | NEC | 0<br>(distal limb in ascending colon) | 1135 | 60 |
| 4. | F | Pacifica | Milk curd obstruction | 25 | 1900 | 120 |
| 5. | M | NZ European | Milk curd obstruction | 60 | 2550 | 177 |
| 6. | F | Māori | NEC | 5 | 2000 | 110 |
| 7. | F | NZ European | NEC | 2 | 3667 | 161 |
| 8. | M | Asian | Meconium ileus cyst | 20 | 1800 | 24 |
| 9. | F | Māori | NEC | 45 | 1250 | 29 |
| 10. | F | Asian | Gastroschisis | 0<br>(distal micro colon) | 2340 | -219 |
| Mean | N/A | N/A | N/A | 21 | 1903 | 68.8 |

### Device and Clinical outcomes

Device and parenteral nutrition (PN) use for each neonate is outlined in **Table 2**. The median device use period was 37.5 (range 12-84) days. CRT was administered through to the morning of enterostomy reversal in all neonates. Chyme was reinfused continuously via the use of a syringe driver, with all chyme that was contained in the stoma bag being reinfused. Specifically, chyme emptied from the proximal limb, into the stoma appliance and was collected every 4 - 6 hours and then reinfused over the following corresponding 4 - 6 hour period. While the reinfusion was occurring, chyme from the proximal limb was able to continue to be collected in the stoma appliance. All chyme was effectively reinfused, with negligible volume loss when a stoma appliance leaked or was replaced.

**Table 2.** Device use and clinical outcomes. *Expressed breastmilk (EBM); **Fortified expressed breast milk (FEBM)

| Neonate no. | Duration of device use (days) | Baseline PN volume (mL/24Hr) | Time to wean from PN (days) | Reduction in PN volume if not weaned (mL/24Hr) | Oral nutrition source | Average weekly weight gain on CRT (g/week) | Average continuous reinfusion rate (mL/Hr) |
| --- | --- | --- | --- | --- | --- | --- | --- |
| 1. | 12 | N/A |  |  | *EBM then Pepti-Junior | 219 | 14 |
| 2. | 31 | 240 |  | No reduction | Neocate | 308 | 8 |
| 3. | 62 | 204 | 14 |  | **FEBM | 179 | 3 |
| 4. | 30 | 582 | 30 |  | *EBM then Pepti-Junior | 292 | 5 |
| 5. | 19 | 90 |  | No reduction | Pepti-Junior | 199 | 7 |
| 6. | 55 | N/A |  |  | Pepti-Junior | 180 | 2.5 |
| 7. | 84 | 550 | 49 |  | Neocate | 40 | 9 |
| 8. | 34 | N/A |  |  | **FEBM | 145 | 3.5 |
| 9. | 48 | 196 |  | Variable | *EBM and Pepti-Junior | 145 | 6 |
| 10. | 41 | 156 | 27 before gut rest | Required gut rest so restarted on PN | *EBM, then Pepti-Junior, then Neocate | 258 | 6 |
| Mean | 37.5 | 204 | 28.5 | N/A | N/A | 197 | 6 |

Neonate discomfort and amount of refluxed chyme flowing back into the stoma appliance was used to judge CRT tolerance, which was variable. Some neonates with a history of feed intolerance were started on a half rate for 24 hours before moving to the full rate, as demonstrated in **Table 2**. Surplus chyme from the neonate receiving chyme reinfusion at a half rate was discarded over that 24-hour period only.

The mean daily weight gain prior to commencing CRT was 68.8 ± 37.4 g/week (individual results per **Table 1**). Following CRT, all neonates had substantial improvements in their daily weight gain, increasing to 197 ± 25.0 g/week (p=0.024) (**Table 2**). Neonatal weight gain before and after CRT are shown in **Figure 4** with examples of weight trajectories before vs after CRT in **Figure 5**. In addition, of 7 neonates dependent on PN at commencement of CRT, 4 were able to wean off, in a median time of 28.5 (range 14-59) days, thereby increasing weight gain while become enterally autonomous with oral feeds and CRT until the time of enterostomy reversal.

**Figure 4.**
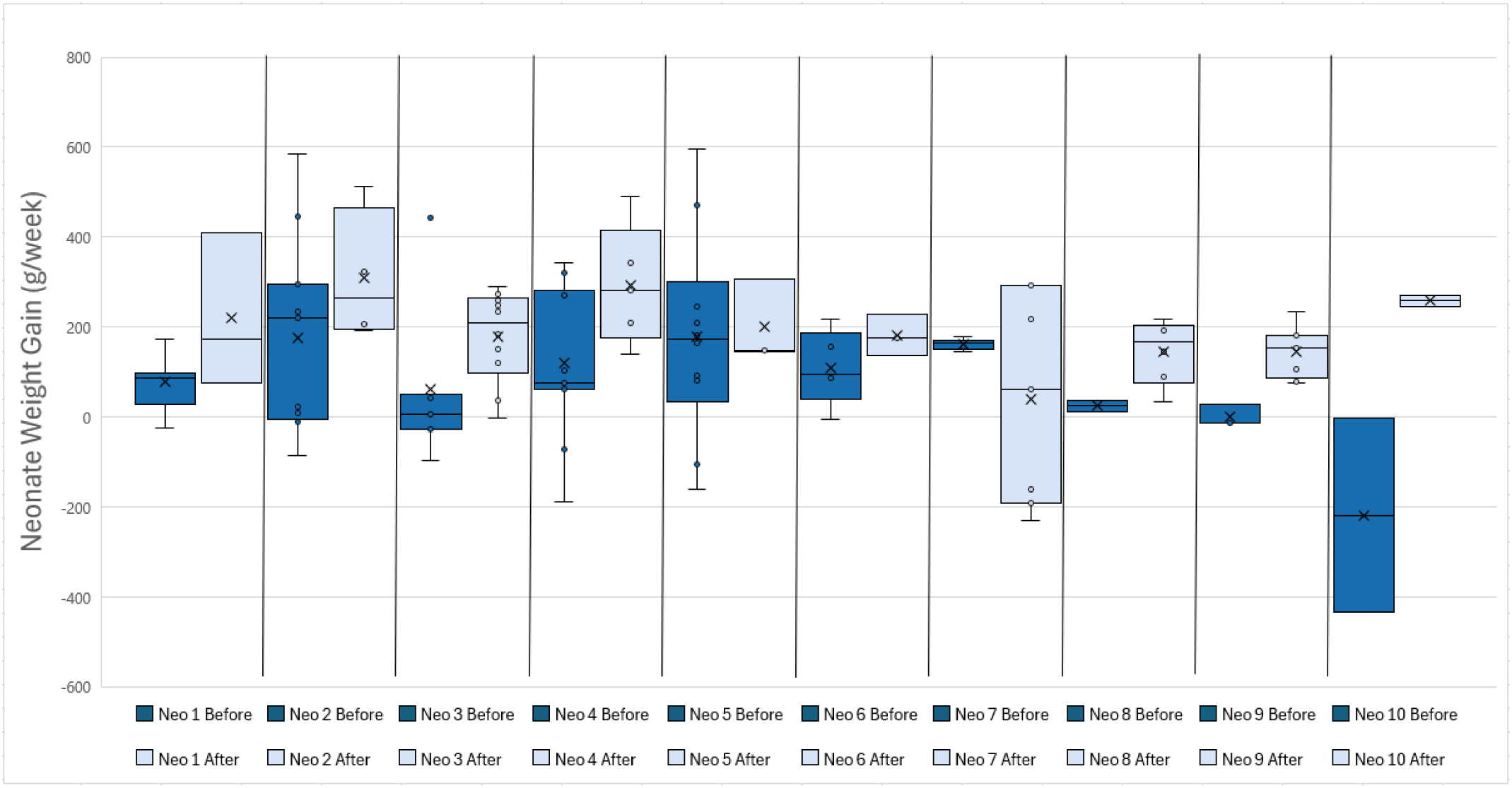
Graph showing neonatal weight gain before and after commencing chyme reinfusion therapy

**Figure 5.**
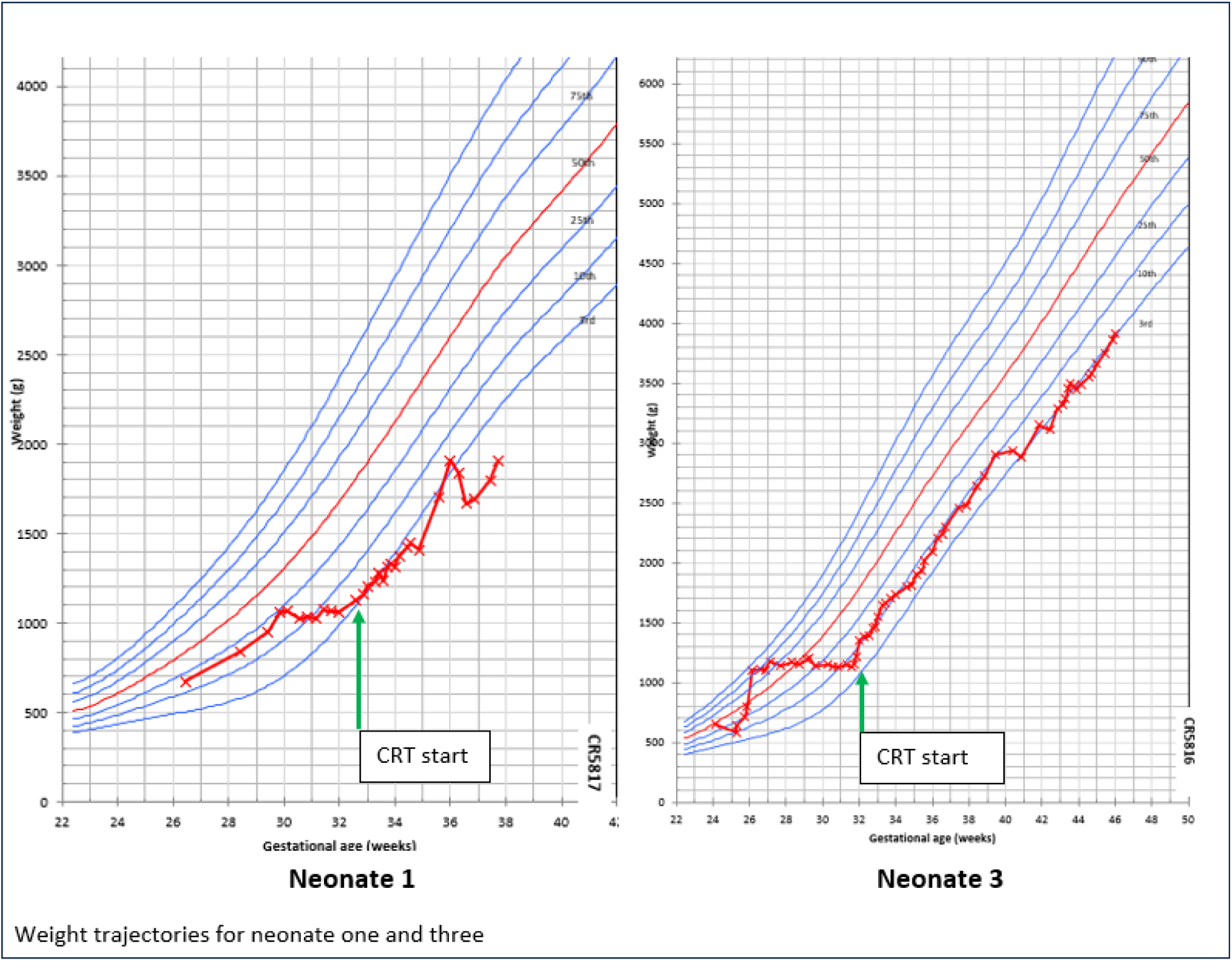
Fenton graphs demonstrating weight trajectories for two neonates

All neonates had their enterostomies reversed, with one suffering a distal limb prolapse that led to an early reversal decision (**Supplementary Table 1**). Although surgical feedback was not included in the protocol, a surgeon provided summary verbal feedback, describing that during enterostomy reversal, the calibre of the proximal and distal limbs matched, enabling easier suturing, stating “The quality of the distal bowel was stunning”.

None of the neonates experienced ileus after reversal surgery. The median time to first bowel movement post-reversal was one day (range 1-3 days). Two patients suffered anastomotic leaks, one of which was managed conservatively, the other requiring reformation of enterostomy. This latter subject had multiple congenital comorbidities that have impacted their recovery, but at the time of writing is being prepared to start CRT again.

### Case Vignettes

Two individual case studies written by a NICU nurse are available upon request from the corresponding author. The case studies demonstrate individual outcomes for the neonates and provide the nursing experience of using the device to administer CRT and manage a neonate with a double enterostomy in intestinal failure.

### Device Usability

#### Stoma appliance use

Overall, the frequency of stoma appliance changes was not impacted by the initiation of CRT. Half of the neonates were reported to have difficult abdominal landscapes, which made keeping a stoma appliance in place and intact challenging. Those neonates continued to have stoma appliance leaks, including occasional peristomal excoriation that is consistent with frequent leaks, which did not change following the initiation of CRT. Comparatively, the other half of the neonates with easier abdominal landscapes were able to maintain wear times of 4 days, their peristomal skin remained healthy, and their stoma appliances and the devices were changed on a schedule.

#### Nursing Usability Assessment

Overall, 10 feedback forms were completed and returned by the cot side nurses with the results outlined in **Table 3**. As demonstrated in **Table 3**, there was a uniformly positive impact on nursing workflow with the device easy to learn and implement. Feedback from cot side nurses, outside of the study feedback form, commented that once they became familiar with the assembly and application of the device, they were astounded by the time saved. With manual reinfusion, the time spent ‘spot repairing’ and manipulation of the stoma appliance on the neonate to prevent leaking was considerable and more than likely, unsuccessful. There was one technical error where a feeding tube was cut prior to being inserted into the distal limb. This required a short period of gut rest out of caution, before recommencing CRT.

**Table 3.** Nursing feedback form results (Responses 1 -Totally disagree, 2 - Disagree, 3 - Neither disagree nor agree, 4 - Agree, 5 - Totally agree)

| Nurse | I believe the neonate stoma refeeding protocol made the refeeding process easier | The number of episodes of reflux of chyme was lower than before | The number of stoma bag changes required was lower than before | I believe the neonate stoma refeeding protocol created a positive impact on workflow | The protocol made the process faster and occurred more smoothly than before | I believe the need for maintenance of the device was low | Pausing chyme reinfusion due to problems arising from the system was less frequent than before | I believe the amount of time needed to become proficient at following the protocol was low | I feel confident to apply the protocol to future patients | I consider the learning process easy to familiarise with for anyone who never applied the protocol |
| --- | --- | --- | --- | --- | --- | --- | --- | --- | --- | --- |
| 1 | 5 | 3 | 4 | 5 | 5 | 4 | 4 | 3 | 5 | 4 |
| 2 | 5 | 4 | 5 | 5 | 5 | 5 | 4 | 4 | 4 | 4 |
| 3 | 5 | 4 | 5 | 5 | 5 | 4 | 3 | 3 | 3 | 4 |
| 4 | 4 | 4 | 2 | 2 | 4 | 4 | 3 | 4 | 4 | 5 |
| 5 | 5 | 4 | 3 | 4 | 5 | 4 | 4 | 5 | 5 | 5 |
| 6 | 5 | 5 | 2 | 5 | 4 | 5 | 3 | 4 | 5 | 5 |
| 7 | 5 | 3 | 3 | 4 | 4 | 3 | 3 | 4 | 4 | 4 |
| 8 | 4 | 3 | 3 | 4 | 4 | 3 | 3 | 4 | 4 | 4 |
| 9 | 4 | 3 | 2 | 4 | 4 | 4 | 3 | 4 | 4 | 4 |
| 10 | 5 | 3 | 3 | 5 | 5 | 5 | 4 | 5 | 5 | 5 |
| Median | 5 | 3 | 3 | 5 | 4-5 | 4 | 3-4 | 4 | 4 | 4 |

### Safety Data

There were no minor or major adverse events attributable to the device. One neonate had unidentified sepsis requiring gut rest during CRT; this neonate returned a positive blood culture of klebsiella with no known source.

## Discussion

This study has introduced a novel device (‘The Insides Neo’) to enable efficient and safe CRT in neonates who have double barrel enterostomy. The device was designed to resolve several limitations of chyme reinfusion identified in previous systematic reviews [11,12], and was manufactured to medical device standards to enable a straightforward and standardized CRT technique. The device was validated within a NICU setting to be safe and effective, and with improved nursing workflows.

The Insides Neo device was designed to fit with off-the-shelf stoma appliances to reduce contact with chyme in comparison to manual CRT, which was found successful in the validation study. Device efficacy was clearly demonstrated by the successful performance of CRT in all patients over a median of more than five weeks, allowing accelerated weight gain and cessation of PN in a majority of subjects. This is a particularly significant outcome of this clinical study, given that PN is accompanied by significant risks including line sepsis, venous thrombosis, and liver impairment, and is an expensive therapy, costing on average $164-239 per bag in US NICU settings, without considering the adjacent costs of PN therapy [15,16]. Therefore, the use of the device in this trial would be expected to lead to cost savings of $USD1,148 - 1,673 / week in weaned patients from PN bags alone, and likely substantial additional savings in treating complications, with a mean cost attributable to blood stream infections secondary to PN of $USD16,141 [16].

The novel device dramatically improved nursing workflows in this study, as evidenced by highly positive responses in related usability domains, as these nurses were able to compare against their prior experiences with manual CRT. Manual CRT typically leads to complexities in modifying stoma appliances and requires a significant time burden, which in return, impacts parental contact with their child [3, 11, 14]. Comparing the assembly of components and equipment between manual CRT and the novel device, the cot side nurse is unable to prepare the components ahead of time with manual CRT due to the frequency of tube installations required to reinfuse and if there is any securement with the stoma appliance, this needs to be modified and completed once in situ [11]. Assembly of the novel device can be completed ahead of time, so when indicated, the replacement and application of the stoma appliance/novel device is a swift process. Goals of reinfusion need to be identified prior to commencing so the correct administration is selected. Both tertiary NICUs in this study chose to administer CRT via continuous reinfusion rather than bolus to reduce reflux which also had a positive impact on nursing time. The benefit of bolus reinfusion is that it mimics peristalsis physiology whereas, continuous reinfusion allows for different feeding tolerances, increasing the rate of absorption due to the slower rate of reinfusion; the novel device allows for varied administration [17].

A further benefit of chyme reinfusion was advanced maturation of the distal (defunctioned) intestine, which enabled improve post-operative outcomes, including rapid return of bowel function (median 1 day) and no cases of ileus. Mismatch of an atrophied or growth-restricted defunctioned distal bowel segment presents a surgical challenge during restoration of continuity [8,14], and anecdotal feedback in this study provided positive indications that CRT can completely resolve this problem. This could be further assessed in future studies by including formal surgical feedback and comparisons of bowel morphology at the time of surgical restoration, and formally assessing surgical complication rates with and without CRT.

Potential CRT-related complications and adverse events highlighted by the Bhat et al., 2020 systematic review were not seen in this study [11], with no device-related morbidity observed. A previous three-arm randomised controlled trial by Lee et al. in 2023, investigating manual CRT on neonates with high output enterostomies, normal output enterostomies, and controls did have a similarly low number of events to this study, while highlighting the inefficiencies around previous CRT administration techniques [14]. These recent studies called attention to the robust weight gain and intestinal adaptation seen with neonates who are distally reinfused, in comparison to non-reinfused neonates, as seen dramatically again in our current data. These findings urge a standardized protocol and administration practices for CRT, which this study has shown can be effectively based on device innovation.

The strength and limitations of this study should be noted. This study introduces a significant new innovation for CRT in neonates and establishes feasibility, with data analysed from the first ten neonates utilising the device. We are now recruiting further patients with the intention to develop a registry and perform further analyses on a larger cohort. This will allow increased statistical power to analyse the benefits and risks associated with using this novel device and inform development of guidelines and protocols for its use. A limitation of this analysis was that the second tertiary site included in this study had not previously performed manual CRT, so nurses from this site did not supply anonymous feedback forms. It would be valuable in future to understand the learning and application process from a novice perspective. It was, however, subjectively noted that there was an easier transition to go from manual CRT to the device at our primary study site, vs all staff starting and learning CRT with the device at the secondary site, likely due to background information and benefits of CRT in general.

In conclusion, this study has presented a novel device ‘The Insides Neo’ to improve nursing workflow when performing CRT in neonates with double enterostomies. The novel device demonstrates safe, effective, and clinically beneficial outcomes in an initial clinical study. The device has now received regulatory approvals in New Zealand, Australia, Europe, and the USA. With increasing awareness and adoption, we anticipate that this innovation will enable the compelling clinical benefits of CRT to become widely available to neonates, while improving the nursing workflows and reducing costs of care.

## Data Availability

All data produced in the present study are available upon reasonable request to the authors

## Acknowledgements

The research team would like to thank the Auckland NICU and Waikato NICU teams who participated in this feasibility study. This paper has been published in a preprint repository and can be found here: www.medrxiv.org/content/10.1101/2024.05.08.24306960v1

## Author Contributions

E. Ludlow: Validation, Formal Analysis, Writing-Original Draft

T. Harrington: Validation, Data Curation

R. Davidson: Conceptualisation, Methodology, Validation, Resources

J. Davidson: Formal Analysis, Resources

K. Aikins: Validation, Writing – Review & Editing

G. O’Grady: Conceptualisation, Methodology, Writing – Review & Editing

1. Bissett: Writing – Review & Editing, Supervision

## Conflict of Interest

E. Ludlow is an employee and shareholder in The Insides Company

T. Harrington contracts to The Insides Company

R. Davidson, J. Davidson, G. O’Grady, and I. Bissett are Founders of The Insides Company

## Funding

The study was funded by a grant from Cure Kids to hire a research nurse and assist with research and development costs associated with The Insides Neo. Devices were supplied by The Insides Company.

## Abbreviations

NEC: Necrotising enterocolitis
CRT: Chyme Reinfusion Therapy
PN: Parenteral Nutrition
EN: Enteral Nutrition
NICU: Neonatal Intensive Care Unit

## Notes

### Clinical Trial

ACTRN12621000835842p

### Author Declarations

New Zealand Health and Disability Committees The Northern B Health and Disability Ethics Committee approved the study 16/Aug/2021 Ethics ref: 21/NTB/150 Study title: Neonate stoma refeeding device feasibility study

### Summary of Updates

This version of the manuscript has been revised and the Discussion and Methods have been updated with more clarification

