## Supplemental Table 1 for "Evaluating the Efficacy and Safety of Neonatal Chyme Reinfusion Therapy: A Feasibility Study using a Novel Medical Device"

| **Neonate no.** | **Stoma Reversed** | **Post -op complications** | **Ileus** | **Time to first BM (days)** |
| --- | --- | --- | --- | --- |
| 1. | Yes | No | No | 1 |
| 2. | Yes | Anastomotic leak not requiring surgical intervention | No | 1 |
| 3. | Yes | No | No | 2 |
| 4. | Yes | No | No | 1 |
| 5. | Yes | No | No | 1 |
| 6. | Yes | No | No | 1 |
| 7. | Yes | No | No | 1 |
| 8. | Yes | No | No | 3 |
| 9. | Yes | No | No | 2 |
| 10. | Yes | Anastomotic leak requiring re-stoma formation. | No | 1 |

**Supplementary Table 1. Clinical outcomes of stoma closure**
